# Prevalence of 406 rare diseases by ethnicity and their associated COVID-19 infection burden: A national cross-sectional study of 62.5 million people in England

**DOI:** 10.64898/2026.01.13.26344068

**Authors:** Qingze Gu, Seyed Alireza Hasheminasab, Marta Pineda-Moncusí, Chimweta Chilala, Johan H Thygesen, Honghan Wu, Sara Khalid, the CVD-COVID-UK/COVID-IMPACT Consortium

**Affiliations:** Centre for Statistics in Medicine, Nuffield Department of Orthopaedics, Rheumatology and Musculoskeletal Sciences (NDORMS), University of Oxford, Oxford, UK; Cardiovascular Epidemiology Unit, University of Cambridge, Cambridge, UK; Institute of Health Informatics, University College London, London, UK

**Keywords:** Rare diseases, Ethnic disparities, Electronic health records, Prevalence, Health inequalities, Population health, COVID-19

## Abstract

**Background:** The COVID-19 pandemic disproportionately affected vulnerable populations, including individuals with rare diseases (RDs) and, in the general population, those from ethnic minority backgrounds. However, the intersectional risk, and how these vulnerabilities combine, is poorly understood. A comprehensive baseline map of RD prevalence by granular ethnicity is required to investigate the pandemic’s true impact on these complex patient groups.

**Methods:** This study had two aims: (1) to generate the national scale prevalence estimates for 406 rare diseases stratified by 19 ethnicity groupings; and (2) to describe the burden of recorded COVID-19 infection across these distinct populations. We conducted a cross-sectional study within the National Health Service (NHS) England Secure Data Environment, accessed via the BHF Data Science Centre’s CVD-COVID-UK/COVID-IMPACT Consortium, linking primary care, hospital, and mortality records for individuals alive on 31 July 2023. We calculated age- and sex-standardised prevalence for 406 RD and calculated COVID-19 infection rates for the overall cohort, for each RD, and by the six-group Office for National Statistics (ONS) and the 19-group NHS ethnicity classifications.

**Results:** We observed a higher burden of COVID-19 infection in the RD cohort (1.7% of the total population) compared to the non-RD population (32.8% vs. 29.0%). The burden varied significantly by ethnicity (e.g., 27.7% in the Pakistani 23.8% in the Black African groups, vs. 33.7% in the White British group) and by specific RD (24.6%–41.2% among the top 25 diseases). This highlights the importance of this ethnicity-stratified prevalence map, which revealed significant underlying ethnic disparities in the RDs themselves. For example, the Pakistani population had markedly elevated odds for certain metabolic disorders (e.g., medium-chain acyl-CoA dehydrogenase deficiency, OR=4.46[4.07–4.90]), and Black Caribbean individuals showed increased odds of autoimmune conditions (e.g., discoid lupus erythematosus, OR=3.34[3.06–3.64]).

**Conclusions:** The burden of the COVID-19 pandemic disproportionately affected individuals with rare diseases. However, the patterning of this risk by ethnicity is complex and runs contrary to general population trends, likely reflecting the deep-seated ethnic disparities in the prevalence of specific RDs. Our foundational map of 406 rare diseases by granular ethnicity is essential for understanding these factors and identifying which specific patient-ethnic subgroups face the greatest intersectional risk.

## Introduction

Though any single rare disease (RD) is uncommon, they collectively pose a significant public health challenge. A RD is defined in Europe as a condition that affects fewer than 1 in 2,000 people [1]. Globally, more than 6,000 known RD affect about 300 million people, or roughly 3.5% to 5.9% of the world’s population [2]. In the UK alone, an estimated 3.5 million people live with an RD [3]. These conditions are often severe and chronic. About 70% have a genetic cause and start in childhood, and nearly a third of affected children die before their fifth birthday [4].

The burden of RD is not limited to its prevalence but extends to substantial diagnostic delays, fragmented care pathways, and significant social and economic impacts for patients and their families. A common feature of RD is the so-called “diagnostic odyssey”[5]—the prolonged and complex journey patients endure to obtain an accurate diagnosis. On average, RD patients experience a delay of 5 years from the onset of symptoms to receiving a confirmed diagnosis[2]. Misdiagnoses are frequent, affecting over 40% of patients, often leading to inappropriate treatments or avoidable complications[6].

While these hurdles affect everyone with an RD, evidence shows that patients from ethnic minority groups often face even greater inequities[7]. Studies report that people from minoritised backgrounds may experience longer diagnostic delays, receive more misdiagnoses, and have less access to specialist care[7]. This is made worse by barriers like language difficulties, cultural misunderstandings, implicit bias, and a historical mistrust of healthcare systems[8, 9]. Additionally, these populations are underrepresented in research, including in genomic databases and clinical trials, which undermines the accuracy of diagnostic tools and genetic tests for these groups[10, 11]. This complex landscape of vulnerability was brought into sharp focus by the COVID-19 pandemic. While prior work has established that the rare disease population faced an increased risk of COVID-19-related mortality and hospitalisation[12], the specific patterning of infection burden by ethnicity within this cohort remains a critical, unanswered question.

Despite the recognised importance of understanding ethnic disparities in RD, national epidemiological data remain limited. Building on our consortium’s recent analysis of this national cohort, which provided prevalence estimates for 331 rare diseases using broad ethnic categories like ‘White’, ‘Asian’, or ‘Black’[12], this study extends that work by applying a more granular framework to an expanded set of 406 conditions. Grouping people by broad categories hides essential differences within these populations. For example, the “Asian” category includes people of Indian, Pakistani, Bangladeshi, and Chinese descent, all with unique genetic and cultural backgrounds that influence their health. These simplified categories can mask differences in disease risk and healthcare access.

Recent advances in data infrastructure now make it possible to analyse ethnicity at a much finer scale. By linking primary care, hospital, and mortality records, the completeness of ethnicity data in the National Health Service (NHS) England Secure Data Environment (SDE) has reached about 94%[13]. Additionally, the development of validated algorithms allows for reconciliation of granular ethnicity coding hierarchies, such as the Systematised Nomenclature of Medicine Clinical Terms (SNOMED CT), with standard Office for National Statistics (ONS) and NHS Digital classifications[13]. This enables analyses at multiple levels of granularity, from six-group ONS categories to 19-group NHS classifications, and down to over 250 self-identified ethnicity subgroups where data volume allows. These advances provide an opportunity to conduct detailed, ethnicity-resolved studies on a national scale.

Improving equity in RD care is also a major policy priority. The UK Rare Diseases Framework[14] and England’s subsequent Action Plans[3] call for reducing inequalities in diagnosis and access to care. These policies emphasise using large-scale health data to find and fix health disparities. At the same time, initiatives like the NHS Genomic Medicine Service and Genomics England’s “Diverse Data” programme aim to ensure that advances in genomics benefit all communities, especially those historically underrepresented in research[15].

In this context, and to address the urgent questions raised by the COVID-19 pandemic, our study had two primary aims. First, to provide the foundational data needed to understand this complex intersection, we aimed to generate the national, population-wide prevalence estimates for 406 clinically curated rare diseases by granular ethnicity. Second, we aimed to apply this new epidemiological map to describe the burden of recorded COVID-19 infection across these distinct rare disease and ethnic populations, providing new insights into how the burden of recorded infection varied across different ethnic groups within the rare disease population.

## Methods

### Study Design, Data Sources, and Population

We conducted a nationwide, cross-sectional study using data in the NHS England SDE, accessed via the BHF Data Science Centre’s <u>CVD-COVID-UK/COVID-IMPACT Consortium</u>. This study leverages a linked, multi-dataset within the NHS England SDE. We used longitudinal primary care data from the General Practice Extraction Service (GPES) Data for Pandemic Planning and Research (GDPPR), secondary care data from Hospital Episode Statistics Admitted Patient Care (HES-APC), Hospital Episode Statistics Outpatient Care (HES-OPC), and mortality records from the Office for National Statistics (ONS) to ascertain rare disease diagnoses and demographics. These were linked to specific datasets used to determine COVID-19 status, including national laboratory COVID-19 testing data from the Public Health England Second Generation Surveillance System, COVID-19 hospital admission data from the COVID-19 Hospitalisations in England Surveillance System, and COVID-19 vaccination status.

Our study cohort was defined as all individuals from the GDPPR dataset alive on the study index date of 31 July 2023. Consistent with the methodology established in our consortium’s previous work [12], all RD diagnoses recorded between 1 January 1990 and the index date were used to identify cases.

### Rare Disease Phenotyping and Ethnicity Harmonisation

As described previously, we adapted a semi-automated phenotyping pipeline to identify a set of clinically specific RD [12]. The established method begins with a comprehensive list of entities in the Orphanet database and selects those with direct mappings to ICD-10 or SNOMED CT terminologies. To ensure high specificity and avoid misclassifying common conditions, mappings labelled “narrow to broad” are excluded. The established pipeline concludes with manual clinician curation to validate code definitions and assess clinical accuracy [12]. While the previous study identified 331 validated RDs, our analysis leverages a broader temporal observation window, allowing an additional 75 conditions for robust clinical validation. This process resulted in our final, expanded set of 406 RD. A patient was classified as having a rare disease if at least one of the corresponding specific ICD-10 or SNOMED-CT codes appeared in their primary or secondary care record during the study period.

A clinician (CC) assigned each of the 406 selected RD to one of three high-level categories based on its primary physiological system: ‘Cardiovascular’ (e.g., congenital pulmonary valvar stenosis), ‘Metabolic’ (e.g., medium-chain acyl-CoA dehydrogenase deficiency), or ‘Other’ (e.g., polymyalgia rheumatica) (**Supplementary data 1**). At the patient level, individuals were assigned a patient-level category. If a patient had multiple RD belonging to the same category, they were assigned that category. If a patient had been diagnosed with RD spanning more than one of these categories (i.e. one cardiovascular and one metabolic), they were classified as ‘Mixed’. This classification was used to describe the baseline characteristics of the rare disease cohort.

Patient-level ethnicity was established using a previously validated hierarchical algorithm that reconciles over 500 data points (e.g. demographic fields, SNOMED-T concepts) from multiple sources to maximise completeness and granularity[13]. The algorithm prioritises the most recent ethnicity record, first from primary care SNOMED CT concepts, then from primary care NHS codes (using the A to Z notation e.g. B Irish white), and finally from secondary care HES data. This process maps individual records to two nested classification schemes: (i) the six-group high-level ethnicity classification used by Office for National Statistics (ONS) (White, Asian or Asian British, Black or Black British, Mixed, Other, Unknown); and (ii) the 19-group NHS primary care classification (White British, White Irish, Any other White background, Indian, Pakistani, Bangladeshi, Chinese, Other Asian, Black African, Black Caribbean, Other Black, White and Caribbean, White and African, White and Asian, Arab, Other Ethnic Group Other Mixed, Unknown).

### Statistical Analysis

Our statistical analysis was structured to address our two primary aims. First, we calculated the prevalence of 406 RDs stratified ethnicity. Second, we described the burden of COVID-19 infections across these specific RD and ethnic cohorts.

#### Aim 1: RD Prevalence and Ethnic Disparity Analysis

We calculated and report point prevalence for the 406 RD defined as the number of individuals with a diagnosis alive on the index date. These crude rates were then adjusted for age and sex using the direct standardisation method, with the 2021 England Census population serving as the reference standard population[16]. We calculated 95% confidence intervals (CI) for the adjusted prevalence rates based on the variance of the standardised rates.

To investigate demographic disparities, we calculated odds ratios (OR) with 95% CI. Each comparison was based on a 2×2 contingency table constructed from the number of affected and unaffected individuals in a comparison group versus a reference group. For gender disparities, the odds of having a given disease in males were compared to those in females, who served as the reference group. Ethnic disparities were analysed at two levels of granularity. First, at the high-level ONS classification level, the odds of disease for each of the ’Asian or Asian British’, ’Black or Black British’, ’Mixed’, and ’Other Ethnic Group’ populations were compared against the ’White’ population as the reference. Second, to explore within-group heterogeneity, a more granular analysis was performed using the 19-group primary care classification, comparing each of the 18 minority sub-communities (e.g., ’Pakistani’, ’Black Caribbean’) against the ’White British’ population, which provided a more specific reference group. 95% CI for the ORs were calculated using the Woolf method on the log-OR scale for all comparisons.

The disparity analyses were restricted to the 227 RDs with 100 or more affected individuals. To quantify the magnitude of ethnic disparities, we calculated OR and 95% CI. Statistical significance was assessed using Pearson’s chi-square test with a Bonferroni-corrected threshold (p<0.00022) to rigorously account for multiple testing across the distinct disease phenotypes.

#### Aim 2: COVID-19 Infection Burden Analysis

To assess the burden of COVID-19 among patients with RD, we linked patient records to national SARS-CoV-2 testing data from the Public Health England Second Generation Surveillance System, COVID-19 hospital admission data from the COVID-19 Hospitalisations in England Surveillance System, COVID-19 vaccination status, and mortality records from the ONS (methods described in previous consortium work [17]). COVID-19 infection status was defined as any documented positive test result (PCR or lateral flow assay) recorded between the start of the pandemic and the study index date (31 July 2023). We calculated the cumulative proportion of individuals with documented infection (expressed as percentages) for: (i) the overall RD cohort compared to the non-RD population; (ii) each of the 406 rare disease phenotypes; and (iii) the six broad ONS and 19 NHS primary care ethnic groups within the RD cohort. These unadjusted descriptive statistics serve to characterise the absolute infection burden observed within these specific subgroups.

### Data Governance and Reproducibility

In adherence with NHS England’s data protection policies, all results intended for publication were subject to disclosure control. Any patient count below 10 was suppressed and displayed as “<10”, and all other counts were rounded to the nearest multiple of five. All data processing and statistical analyses were performed within the NHS England SDE using Python in Databricks and R 4.2.3, and according to a pre-specified analysis plan published on GitHub, along with the phenotyping code lists, and data curation and analysis code (https://github.com/BHFDSC/CCU069_01).

## Results

### Cohort Characteristics

The study cohort included 62,539,740 individuals, of whom 1,051,470 (1.7%) were identified as having at least one of the 406 curated RD. Baseline demographic characteristics of the rare disease and non-rare disease cohorts are presented in **Table 1**. **Tables S1–3** showed baseline characteristics of the study cohort by rare disease categories, by the six broad ONS ethnicity groups, and by the nineteen NHS primary care ethnicity categories. Overall, individuals in the rare disease cohort were more likely to be female than in the non-rare disease cohort (54.7% vs. 49.9%), and had a slightly older median age at index date (46 [IQR 26–79] vs 40 [23–58] years). The overall ethnic distribution showed a higher proportion of White individuals in the rare disease cohort. Within the rare disease cohort, most patients only had one rare disease (median 1, IQR 1–1), and the majority of patients were classified into the “Other” disease category, with a smaller but substantial proportion having Cardiovascular, Metabolic, or Mixed-category diseases.

**Table 1.**
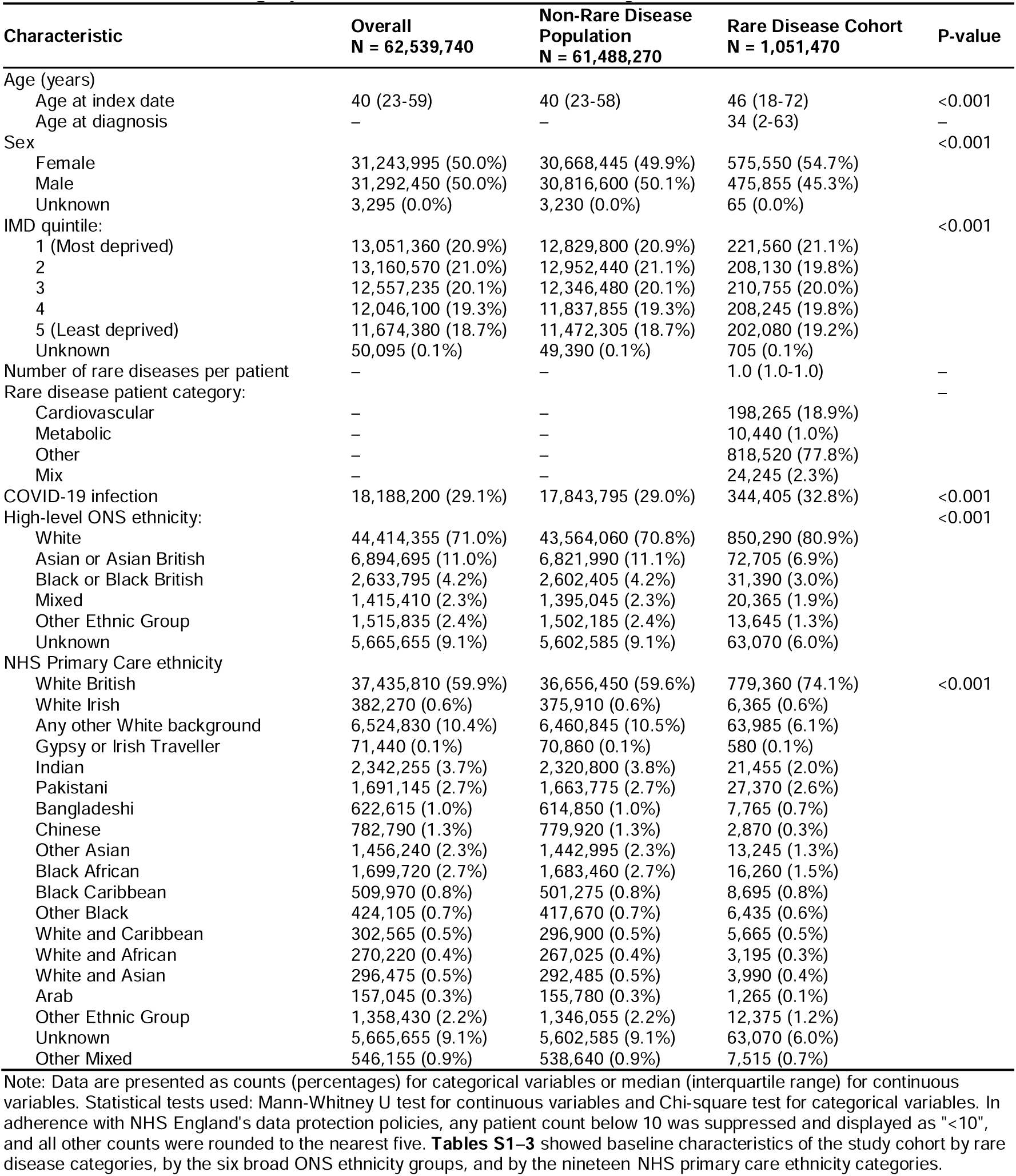
Baseline Demographic Characteristics of the Study Cohort.

### Prevalence and Disparities in Rare Diseases

The age- and sex-standardised prevalence of the 406 RD varied substantially (detailed prevalence estimates in Supplementary data 2). **Figure 1A** presents the 25 most common RDs. The most prevalent conditions were polymyalgia rheumatica (3,139.5 [95% CI: 3,124.9–3,154.1] per million), interatrial communication (2,449.7 [2,436.8–2,462.7] per million), and infantile apnoea (1,057.5 [1,049.0–1,066.0] per million).

**Figure 1.**
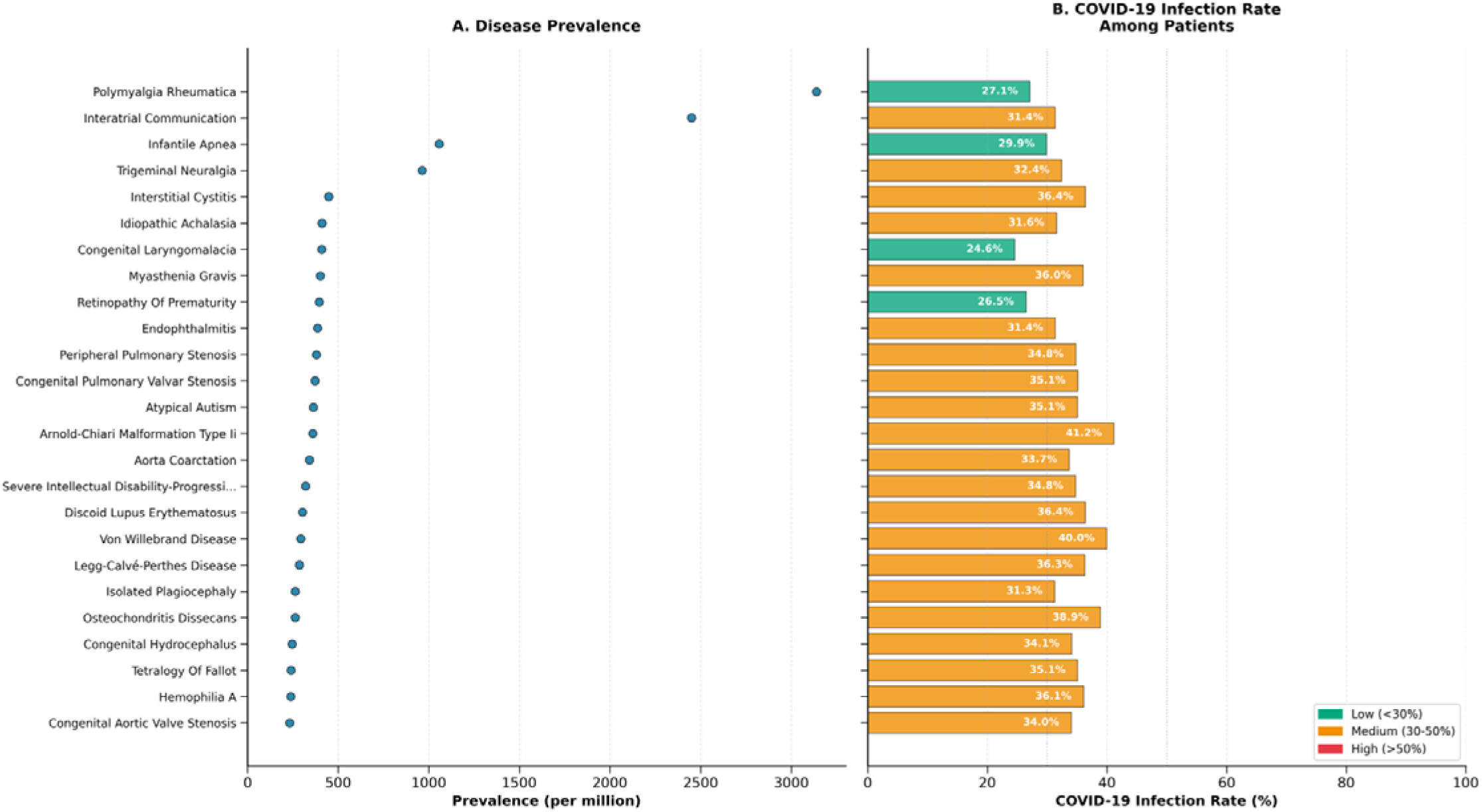
Prevalence and COVID-19 Infection Burden of the Top 25 Rare Diseases. Forest plot showing age- and sex-standardised prevalence (A) and COVID-19 infection rates (B) for the 25 most common rare diseases among 62.5 million individuals in England (study end date: 31 July 2023). Panel A: Prevalence per million population, standardised to the 2021 England Census population. Panel B: Percentage of patients with documented SARS-CoV-2 infection during the pandemic period. Colours indicate COVID-19 burden categories: green = low (<30%), yellow = medium (30–50%), red = high (>50%). Vertical reference lines at 30% and 50% aid interpretation. Four diseases showed low infection rates: polymyalgia rheumatica (27.1%), infantile apnoea (29.9%), congenital laryngomalacia (24.6%), and retinopathy of prematurity (26.5%). All other diseases demonstrated medium rates (30–50%). Diseases are ranked by prevalence (Panel A) and displayed in the same order across both panels for direct comparison.

#### Prevalence by Ethnicity

The prevalence of the ten most common RDs also differed substantially across broad and detailed ethnic categories, as shown in the prevalence heatmaps (Figure 2, detailed estimates in Supplementary data 3). When using the six high-level ONS ethnic groups (**Figure 2A**), Polymyalgia Rheumatica exhibited the highest prevalence in the White group (3,696.8 [3678.9–3714.7] per million), with lower rates observed in Asian or Asian British (456.7 [440.8–472.7] per million) and Black or Black British (403.9 [379.7–428.3] per million) categories. Interatrial Communication was most prevalent in the Mixed group (3,006.2 [2915.9–3096.5] per million), followed by White (2,331.5 [2317.3–2345.7] per million). Further analysis using nineteen detailed NHS primary care ethnicity categories revealed additional heterogeneity (**Figure 2B**). For instance, the prevalence of Interatrial Communication was highest in the Mixed White and Caribbean group (3,721.5 [3504.2–3938.9] per million), which was 1.6 times greater than in the Mixed White and African group (2,372.1 [2188.5–2555.8] per million).

**Figure 2.**
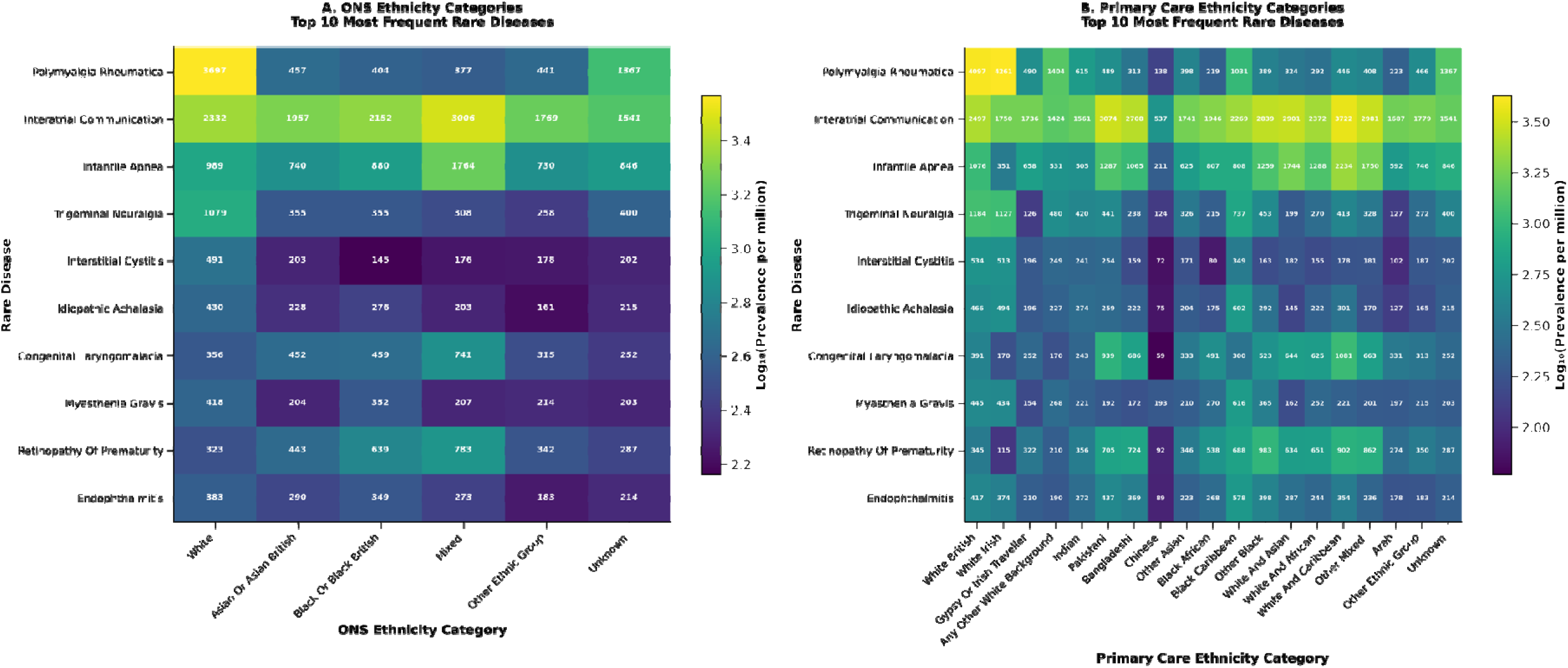
Prevalence of the Top 10 Most Frequent Rare Diseases by Ethnicity: ONS High-level and NHS Primary Care Classifications. Side-by-side heatmaps displaying the prevalence (per million) of the ten most frequent rare diseases across major ethnic groups, using (A) 6 high-level ONS ethnicity categories and (B) 19 NHS primary care ethnicity categories. Prevalence values are coloured on a logϑϑ scale for visual clarity, with absolute values annotated within each cell.

#### Ethnic Disparities in Rare Disease Risk

A summary of the odds ratios for all 227 diseases across the high-level ONS ethnicity groups shows widespread and significant disparities (**Figure S1**). Further investigation at both the high-level ONS and granular primary care levels revealed distinct risk patterns (detailed estimates in Supplementary data 4–5). At the ONS level, 104 diseases showed significantly different prevalence in Asian or Asian British individuals compared to White individuals (38 higher, 66 lower). Notable elevations included Lamellar ichthyosis (OR 5.75 [95% CI 4.48–7.38]) and Maple syrup urine disease (OR 5.71 [4.21–7.74]), while conditions like Polymyalgia rheumatica showed significantly lower prevalence (OR 0.12 [0.12–0.13]). Similarly, 83 diseases differed significantly for Black or Black British individuals (28 higher, 55 lower), with higher risks observed for Moyamoya disease (OR 4.47 [3.71–5.37]) and lower risks for conditions such as Porphyria cutanea tarda (OR 0.08 [0.03–0.21]). For the Mixed ethnicity group, 54 diseases showed significant disparities (32 higher, 22 lower), including elevated risks for Isolated congenital sclerocornea (OR 3.53 [2.35–5.30]).”

#### Heterogeneity within Asian or Asian British Groups

Forest plots were generated for each ethnic subgroup to visualise ethnic disparities in RD risk. These plots display up to 20 diseases with the highest patient counts within that subgroup, ordered by their odds ratio, to highlight the most prevalent and significant associations. At the high-level ONS level, individuals of Asian or Asian British ethnicity exhibited the highest odds for Kawasaki disease (odds ratio (OR)=1.55, [95% CI 1.46–1.64]), retinopathy of prematurity (OR=1.37 [1.32–1.43]) and congenital laryngomalacia (OR=1.27 [1.22–1.32]) compared to White individuals (**Figure 3A**).

**Figure 3.**
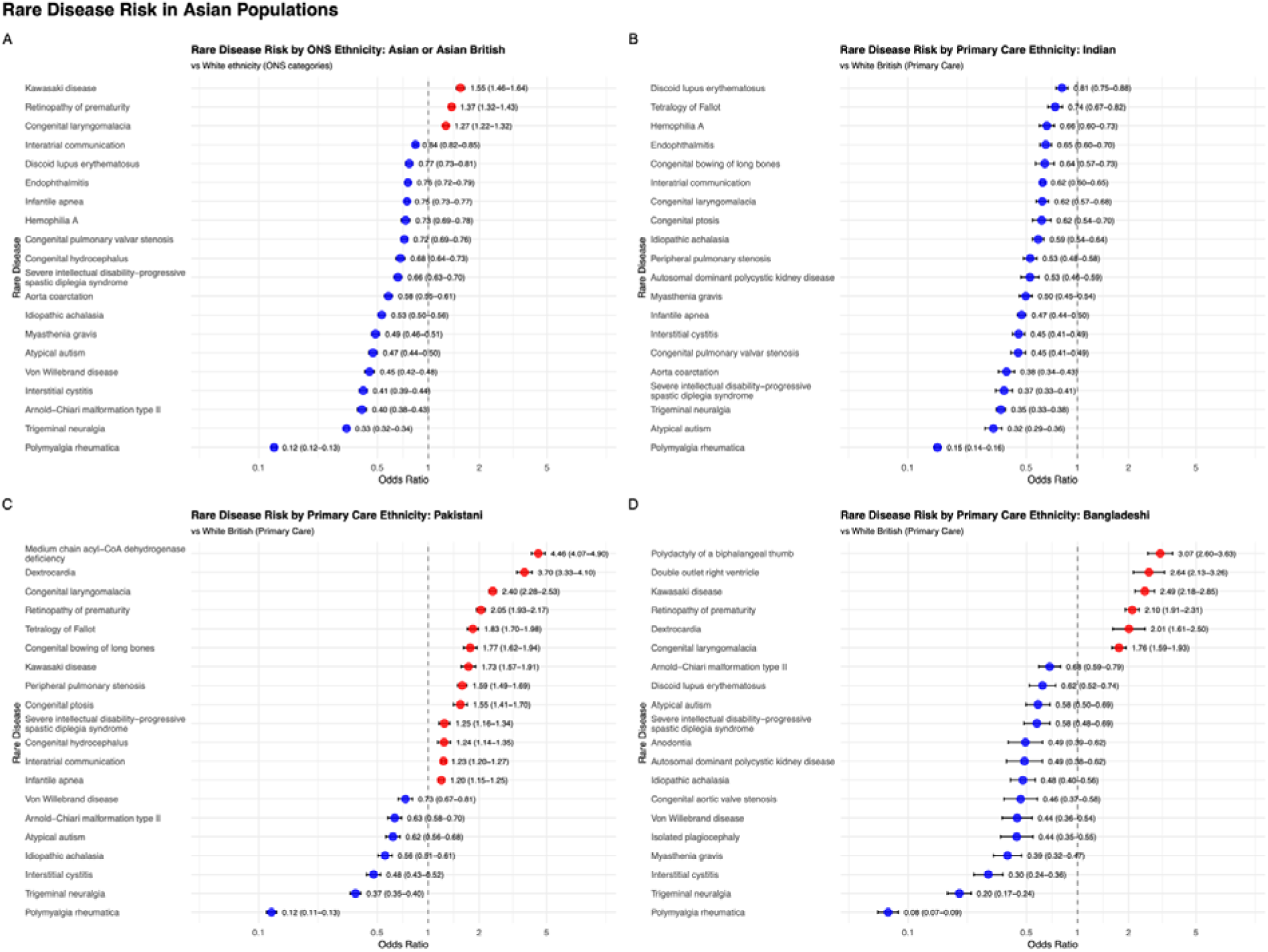
Rare Disease Risk in Asian Populations. Four-panel forest plots displaying the top 20 rare diseases with the highest patient counts and odds ratios in Asian populations, compared to White or White British reference groups. Panel A shows results for the broad ONS “Asian or Asian British” category (vs White ethnicity). Panels B, C, and D present results for the Indian, Pakistani, and Bangladeshi subgroups, respectively, using NHS primary care ethnicity classifications (each vs White British). Odds ratios (OR) and 95% confidence intervals are shown for each disease.

However, analysis using the 19-group primary care data revealed that these risks were not uniform across all subgroups and were primarily driven by specific ones (**Figure 3B-D**). The Pakistani population had a much higher risk for medium chain acyl-CoA dehydrogenase deficiency (OR=4.46 [4.07–4.90]), dextrocardia (OR=3.7 [3.33–4.10]), congenital laryngomalacia (OR=2.40 [2.28–2.53]), and retinopathy of Prematurity (OR=2.05 [1.93–2.17]) compared to White British individuals (**Figure 3C**).

The Bangladeshi community also showed elevated odds for Kawasaki disease (OR=2.49 [2.18–2.85]), retinopathy of prematurity (OR=2.10 [1.91–2.31]) and congenital laryngomalacia (OR=1.76 [1.59–1.93]). Additionally, this group demonstrated higher risks for specific conditions than British White individuals, including polydactyly of a biphalangeal thumb, double outlet right ventricle, and dextrocardia (**Figure 3D**). Conversely, for many conditions, individuals of Indian ethnicity showed a lower risk compared to the White British population (**Figure 3B**).

#### Heterogeneity within Black or Black British Groups

A similar pattern of heterogeneity was observed within Black or Black British subgroups (**Figure 4**). While the aggregate ONS group showed the highest risk for Kawasaki disease (OR=2.33 [2.17–2.50]), congenital subglottic stenosis (OR=2.27 [2.05–2.51]), and retinopathy of prematurity (OR=1.98 [1.88–2.08]) (**Figure 4A**) compared to White individuals, the granular data pinpointed more specific and potent risk profiles. The Black Caribbean population exhibited exceptionally high odds of discoid lupus erythematosus (OR=3.34 [3.06–3.64]), lichen planopilaris (OR=3.18 [2.41–4.19]), neuromyelitis optica spectrum disorder (OR=3.08 [2.43–3.92]), and idiopathic avascular necrosis (OR=3.07 [2.44–3.87]) compared to White British individuals (**Figure 4C**). The Other Black group displayed the highest odds for Kawasaki disease (OR=3.09 [2.68–3.57]) and a more pronounced risk for retinopathy of prematurity (OR=2.85 [2.50-3.14]) than the aggregate Black group (**Figure 4D**). Individuals of Black African ethnicity also had elevated risks for congenital subglottic stenosis (OR=1.98 [1.75–2.25]), Kawasaki disease (OR=1.86 [1.69–2.05]), and retinopathy of prematurity (OR=1.56 [1.46-1.67]). However, it was less pronounced than in other Black subgroups (**Figure 4B**).

**Figure 4.**
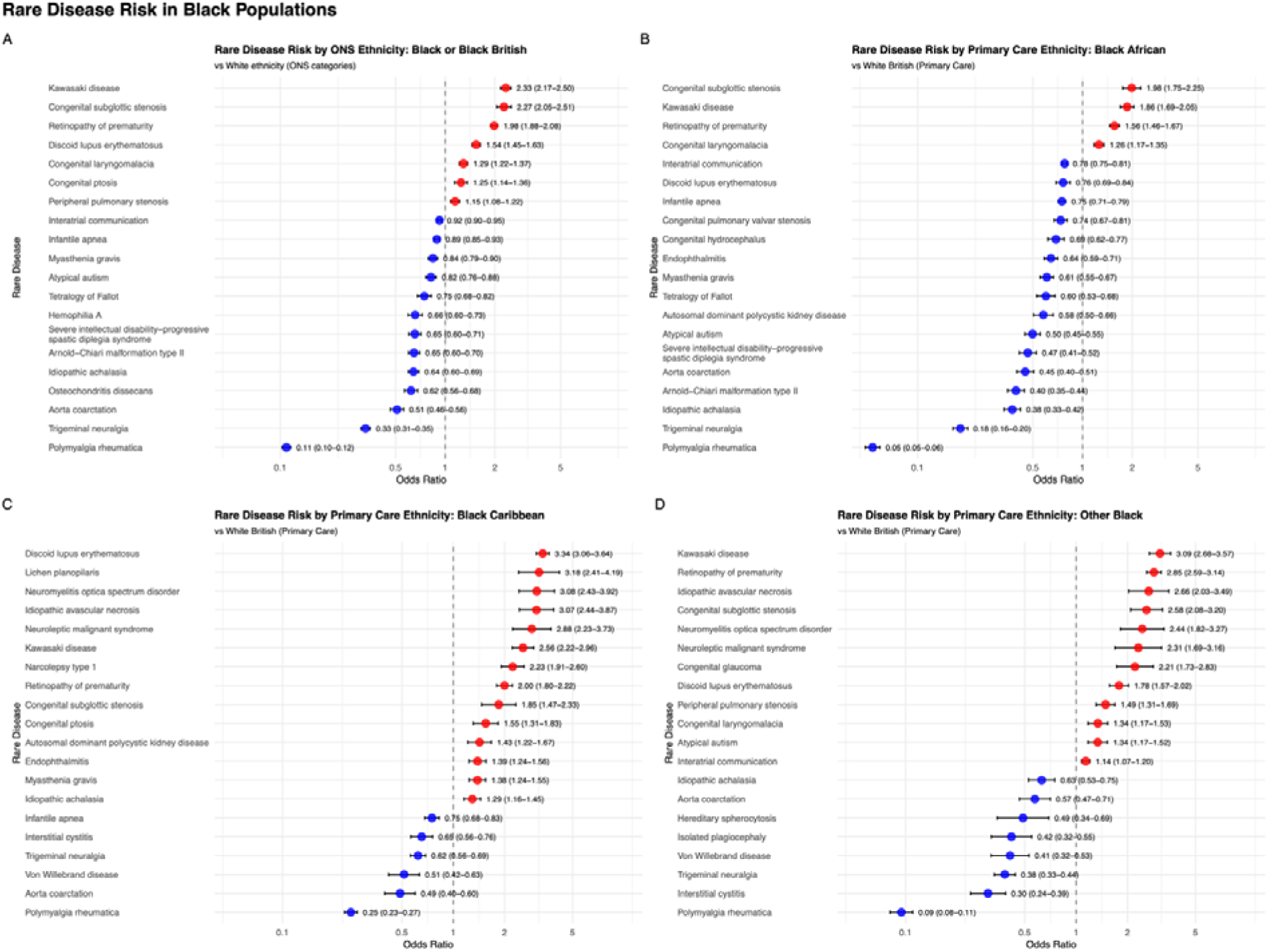
Rare Disease Risk in Black Populations. Four-panel forest plots displaying the top 20 rare diseases with the highest patient counts and odds ratios in Black populations, compared to White or White British reference groups. Panel A shows results for the broad ONS “Black or Black British” category (vs White ethnicity). Panels B, C, and D present results for the Black African, Black Caribbean, and Other Black subgroups, respectively, using NHS primary care ethnicity classifications (each vs White British). Odds ratios (OR) and 95% confidence intervals are shown for each disease.

#### Disparities in Mixed Ethnicity Groups

Significant disparities were also identified in individuals of Mixed ethnicity (**Figure 5**). Kawasaki disease demonstrated a consistently elevated risk across all analysed mixed-heritage groups. The OR for Kawasaki disease was 2.70 [2.47–2.95] in Mixed versus White individuals. Within specific subgroups and compared to White British, the OR was 2.91 [2.45–3.47] for individuals of White and Caribbean heritage, 1.95 [1.56–2.44] for White and African, and 2.61 [2.16–3.14] for White and Asian (**Figure 5A–D**). Granular analysis further revealed that certain subgroups were disproportionately affected by specific diseases. For example, individuals of White and Caribbean were overrepresented in cases of congenital laryngomalacia (OR=2.77 [2.48–3.09] compared to White British), those of White and African in congenital subglottic stenosis (OR=2.64 [2.03–3.44]), and those of White and Asian in levocardia (OR=2.69 [1.75–4.15]).

**Figure 5.**
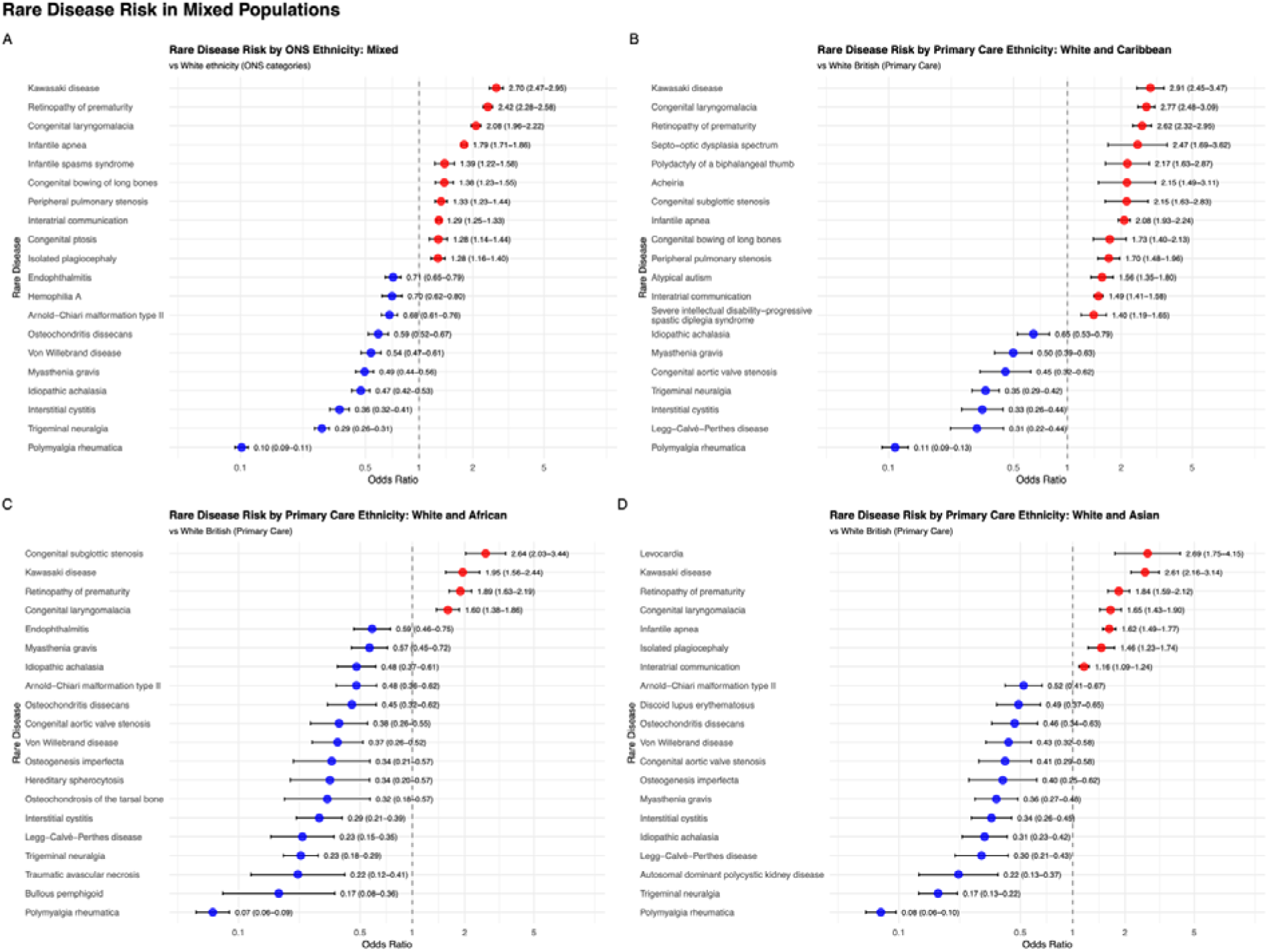
Rare Disease Risk in Mixed Populations. Four-panel forest plots displaying the top 20 rare diseases with the highest patient counts and odds ratios in Mixed populations, compared to White or White British reference groups. Panel A shows results for the broad ONS “Mixed” category (vs White ethnicity). Panels B, C, and D present results for the White and Caribbean, White and African, and White and Asian, respectively, using NHS primary care ethnicity classifications (each vs White British). Odds ratios (OR) and 95% confidence intervals are shown for each disease.

#### Gender-Based Disparities in Disease Risk

Disparities in disease risk between sexes were evident for numerous conditions (detailed in Supplementary data 6). A strong male predisposition was observed for several conditions, excluding those inherently male-specific, such as 47, XYY syndrome and testicular regression syndrome. Notably, males had higher odds for Legg-Calvé-Perthes disease (OR=2.49 [2.41–2.58] compared to females), atypical autism (OR=2.12 [2.06–2.19]), and isolated plagiocephaly (OR=2.10 [2.03–2.18]).

Conversely, a pronounced female predisposition was characteristic of several autoimmune and inflammatory conditions. For example, the odds ratio for males was 0.47 [0.47–0.47] for polymyalgia rheumatica. The odds of interstitial cystitis were over eight times higher in females compared to males (OR=for males 0.12 [0.12–0.13]), and the odds of discoid lupus erythematosus were over five times higher in females (OR=for males 0.19 [0.19–0.20]).

### COVID-19 Infection Burden

We observed a 2.8% higher overall rate of recorded COVID-19 infections in the rare disease cohort compared to the non-rare disease cohort (32.8% vs 29.0%, respectively; Table 1). However, the burden was not uniform across ethnic groups within the RD cohort. Counter to trends in the general population, recorded COVID-19 infection rates were highest in the White British group (33.7%) and the White ONS group (33.5%). These were higher than rates observed in the Black African (23.8%), Gypsy or Irish Traveller (24.1%), Bangladeshi (27.2%), and Pakistani (27.7%) groups (Table S2–3). The COVID-19 infection burden also varied substantially by the specific rare disease. Among the 25 most prevalent RDs, infection rates ranged from 24.6% to 41.2% (**Figure 1B**, rates for all 406 RDs in Supplementary data 7). Four diseases demonstrated relatively lower infection rates (<30%): congenital laryngomalacia (24.6%), retinopathy of prematurity (26.5%), polymyalgia rheumatica (27.1%), and infantile apnoea (29.9%). The remaining 21 diseases all showed medium infection rates (30–50%, shown in yellow).

## Discussion

In this nationwide, cross-sectional analysis of linked EHRs for 62.5 million people in England, we mapped the prevalence of 406 clinically curated RD by ethnic group. We observed that 1.7% of the population had at least one of the studied RD diagnosis and that age- and sex-standardised prevalence varies widely across both six high-level ONS categories and the 19 NHS primary-care ethnicity groups. When we broke ethnicity into finer groups, we saw substantial differences within broad categories that the analysis of the more specific groups uncovered. These findings extend our consortium’s earlier national estimates across 331 conditions, which established sex and broad-ethnicity differences[12], by systematically investigating granular ethnic subgroups at scale. This current study demonstrates the added value of a high-granularity ethnicity resource when applied to RD epidemiology[12, 13].

Our analysis revealed that patients with RD experienced a moderately elevated COVID-19 infection rate (32.8%) compared to the general population without RD (29.0%), consistent with reports of increased vulnerability among individuals with chronic conditions during the pandemic[18]. However, the burden was not uniform across disease phenotypes. Among the 25 most prevalent RD, infection rates ranged from 24.6% to 41.2%, with the majority (21/25) recording rates between 30% and 50%. Notably, the three conditions with the lowest COVID-19 rates—retinopathy of prematurity, congenital laryngomalacia, and infantile apnoea—predominantly affect neonatal and paediatric populations, consistent with lower paediatric infection rates observed during the pandemic. The lower rate observed in polymyalgia rheumatica, which primarily affects older adults, may reflect the success of shielding programmes and vaccine prioritisation for clinically vulnerable populations in England, or potentially survival bias if patients with severe COVID-19 outcomes were excluded from later follow-up.

Our prevalence findings align with and expand upon previous national estimates. For instance, For instance, the three most common RDs, polymyalgia rheumatica, interatrial communication, and infantile apnoea, were the same as those identified in the earlier nationwide analysis[12]. We consistently observed slightly higher prevalence estimates for these conditions (e.g., 3,139.5 vs. 2,831.75 per million for polymyalgia rheumatica), which is likely attributable to our study’s longer follow-up period to July 2023, allowing for more complete case ascertainment. While our consortium’s previous work benchmarked these aggregate figures against Orphanet[12], this study advances that evidence by resolving prevalence for specific ethnic groups. Furthermore, our findings on demographic disparities corroborate the previous work, reinforcing the robustness of these patterns. We confirmed the strong female predominance in autoimmune conditions like discoid lupus erythematosus (male vs. female OR=0.19) and polymyalgia rheumatica (OR=0.47), consistent with prior findings. Similarly, the markedly higher odds of discoid lupus erythematosus in the Black population and the lower odds of polymyalgia rheumatica in both Asian and Black populations, relative to the White population, were consistent across both studies.

Our study adds significant new insight, however, in dissecting these broad categories. Refining the analysis from six ONS groups to the 19 NHS primary care categories changed the interpretation of who is most affected. For example, the higher prevalence of interatrial communication in the ONS “Mixed” category was primarily driven by the “White and Caribbean” subgroup, not uniformly by all mixed-heritage communities. Likewise, conditions with low overall frequency but strong ethnic enrichment emerged only after disaggregation, such as lamellar ichthyosis, maple syrup urine disease, and Wolfram syndrome in the Pakistani population, and neuromyelitis optica spectrum disorder in Black Caribbean individuals. These examples illustrate how analyses that fully explore ethnic heterogeneity can surface inequities relevant to screening pathways, clinical suspicion, and genetic evaluation that would otherwise remain hidden[13].

A central finding of our study was the complex and counter-intuitive patterning of recorded COVID-19 infection by ethnicity within the RD cohort. While ethnic minority groups faced a higher burden of COVID-19 in the general population[18–21], we observed the opposite trend among those with rare diseases. Recorded infection rates were highest in the White British group (33.7%) and notably lower in groups such as Black African (23.8%), Bangladeshi (27.2%), and Pakistani (27.7%). This highlights that vulnerability is not uniform and implies that the specific disease composition of each ethnic cohort drives recorded risk. For example, interstitial cystitis, which is significantly more prevalent in White populations (OR 0.41 in Asian vs White), had a high recorded infection rate of 36.4%. In contrast, retinopathy of prematurity, which showed elevated prevalence in Black (OR 1.98) and Asian (OR 1.37) groups, had a much lower infection rate of 26.5%, likely reflecting the protective shielding of neonatal populations. Thus, the aggregate ethnic disparities observed may be a proxy for the distinct age profiles and shielding behaviours associated with the specific rare diseases clustered within those groups, alongside potential differences in symptomatic testing thresholds.

This intersection of our two aims offers a potential explanation for these complex findings. The differences in rare disease prevalence across ethnic groups likely stem from a complex mix of biological, social, and health-system factors. Genetics may explain some variations; for example, certain conditions are more common in specific communities due to founder effects or a higher prevalence of specific pathogenic variants.[2, 6, 7, 9, 22]. At the same time, many ethnic minority communities face significant barriers that can lead to underdiagnosis and care inequities. Obstacles such as language barriers, lower health literacy, lack of trust in the healthcare system, and fewer or later referrals to specialist services can lengthen the “diagnostic odyssey” and increase the chance of misdiagnosis[9]. These gaps are widened by the historic under-representation of non-European ancestries in genomics and clinical studies. This makes reference databases and risk tools less accurate for these populations, often resulting in a lower diagnostic yield[10, 11]. This creates a crucial challenge for interpreting our results. A lower recorded prevalence for a condition in an ethnic minority group could reflect an actual lower biological risk due to genetic or environmental factors. However, it could also signal systemic under-recognition, where unequal access to care leads to fewer diagnoses.

This means some of our findings may be underestimating the actual disease burden in these communities, potentially masking even greater health disparities. Our results, therefore, point to tangible opportunities for refining diagnostic strategies, such as informing targeted cascade testing and newborn screening pilots[23], and for prioritising the development of more inclusive genomic reference panels[10, 11, 13–15].

This study demonstrates the methodological value of combining RD phenotyping at the population level with a validated, hierarchical approach to ethnicity reconciliation across multiple data sources[12, 13]. By prioritising recent primary-care SNOMED CT concepts, then NHS codes, and finally hospital records, and structuring outputs to both ONS and NHS schemes, we achieved high completeness while retaining granularity for analysis[13]. This approach and routine age- and sex-standardisation provide a pragmatic framework for national RD surveillance and equity audits in other disease areas that rely on EHRs.

### Implications for policy and care

These prevalence and disparity findings have several implications. Clinically, services can use them to refine referral thresholds, triage investigations, and tailor patient-facing guidance for communities at higher risk of specific RD. The systematic sex differences we confirmed suggest that sex-aware diagnostic pathways and trial eligibility criteria may improve both time-to-diagnosis and treatment generalisability. Health-system planners can better anticipate demand for specialist diagnostics and therapies, aligning with the UK Rare Diseases Framework and England Action Plans that call for earlier, fairer diagnosis and improved care coordination[3, 14]. Research sponsors and genomic initiatives can use the highlighted gaps to prioritise recruitment and variant curation in underrepresented groups, supporting equitable benefits from genomic medicine[10, 11]. Finally, analysts should avoid defaulting to five or six ethnicity categories when data permit a more nuanced representation; the present findings show that important signals sit within the “average” of broad groups and become visible only with finer mappings[13].

### Strengths and limitations

Our study has several strengths, including near-complete national coverage of patients registered in a general practice with linkage of primary care, hospital, vaccination, and mortality data; and use of an established workflow to curate clinically specific RD code lists[12, 13]. Building on our consortium’s foundational analysis of 331 rare diseases using broad ethnic categories [12], this study extends that work by providing the first prevalence estimates across fine-grained ethnic categories for an expanded set of 406 conditions, many of which lack contemporary data in traditional registries.

Limitations should be acknowledged. This was a cross-sectional prevalence study; we did not interrogate incidence, diagnostic delay, survival, or causal mechanisms. As with all EHR studies, misclassification is possible due to coding errors, changes in terminology over time, and variable capture across care settings; although we excluded broad mappings and undertook clinical curation, some degree of misclassification may remain[12]. Ethnicity recording, with completeness reaching approximately 94% after linkage, can still be missing, inconsistent, or mapped differently depending on legacy systems; specific categories (e.g., Chinese, Gypsy/Irish Traveller) have differed between census and NHS schemes over time[13]. These issues can bias both numerator and denominator definitions for disparity estimates. Although we adjusted prevalence by age and sex and used conservative multiple-testing thresholds, unmeasured confounding (e.g., migration history, language, and healthcare utilisation patterns) could contribute to observed differences. COVID-19 infection rates in our study are likely underestimated, as they rely exclusively on documented testing data and do not capture asymptomatic infections, untested symptomatic cases, or infections occurring in settings without routine testing. Testing availability and policies varied substantially throughout the pandemic period (January 2020–July 2023). Most notably, the cessation of the universal free testing offer in England on 1 April 2022 markedly reduced data coverage for the final 15 months of the study period [24]. Consequently, ascertainment in the latter stages is heavily skewed towards hospitalised cases and specific clinically vulnerable groups eligible for continued testing, potentially introducing differential bias across rare disease phenotypes. Finally, we did not evaluate how ascertainment, care-seeking, or diagnostic access varied by ethnicity; differential access could amplify or mask true biological differences.

Future work would benefit from focusing on the following aspects. First, longitudinal analyses are needed to quantify diagnostic delay, multimorbidity trajectories, mortality, and healthcare utilisation across ethnic and sex strata, leveraging the already-established linked datasets. Second, integration with genomic resources, including variant databases and sequencing programmes, can test whether the disparities observed here align with allele frequency differences, founder events, or gene–environment interactions, and can inform variant interpretation in historically under-represented populations[10, 15]. Third, methods development should continue for robust, privacy-preserving analytics in granular strata, including hierarchical modelling that shares information across related subgroups and harmonisation with evolving ONS categories to minimise misclassification[13]. Aligning these advances with priorities in the UK Rare Diseases Framework and Action Plans will support translation into screening policies, clinical pathways, and equitable trial designs[3, 14].

In conclusion, national-scale, ethnicity-resolved rare disease epidemiology is feasible and informative. Using a high-granularity ethnicity framework within linked EHRs, we identified invisible inequities in coarse aggregations and provided actionable evidence to support earlier diagnosis, inclusive medications, and fair allocation of specialist services. These results provide a practical foundation for precision public health in RD and a template for equity-focused analyses across other conditions.

## Ethical approval

The North East – Newcastle and North Tyneside 2 research ethics committee provided ethical approval for the CVD-COVID-UK/COVID-IMPACT research programme (REC no: 20/NE/0161) to access, within secure, trusted research environments, unconsented, whole-population, anonymised data from electronic health records collected as part of patients’ routine healthcare. Our project (proposal CCU069, short title: RARE-CVD-COVID) agreed to the objectives of the consortium’s ethical and regulatory approvals and was authorised by the BHF Data Science Centre’s Approvals and Oversight Board. Approved researchers (QG, ASH, MPM, SK) conducted the analyses within NHS England’s SDE via secure remote access. Ensuring the anonymity of individuals, only summarised-aggregated results manually reviewed by the NHS England ‘safe outputs’ escrow service were exported from the SDE.

## Data availability

The data used in this study are available in NHS England’s Secure Data Environment (SDE) service for England (https://digital.nhs.uk/services/secure-data-environment-service). The CVD-COVID-UK/COVID-IMPACT programme led by the BHF Data Science Centre (https://bhfdatasciencecentre.org/) received approval to access data in NHS England’s SDE service for England from the Advisory Group for Data (AGD) (https://digital.nhs.uk/about-nhs-digital/corporate-information-and-documents/advisory-group-for-data) – formerly the Independent Group Advising on the Release of Data (IGARD) – via an application made in the Data Access Request Service (DARS) Online system (ref. DARS-NIC-381078-Y9C5K) (https://digital.nhs.uk/services/data-access-request-service-dars/dars-products-and-services). The CVD-COVID-UK/COVID-IMPACT Approvals & Oversight Board (https://bhfdatasciencecentre.org/areas/cvd-covid-uk-covid-impact/) subsequently approved this project (CCU069) to access the data within NHS England’s SDE service for England. The anonymised data used in this study were made available to accredited researchers. Those wishing to gain access to the data should contact in the first instance.

## Author contributions

The study was designed and conceived by SK and QG. SAH extracted the dataset, QG, SAH, MP, JHT, HW, and CC curated the data. QG created the visualisations. QG analysed the data and wrote the first draft of the manuscript. All authors contributed to interpreting the data and revising the manuscript.

## Acknowledgements

This work was carried out with the support of the BHF Data Science Centre led by HDR UK (BHF Grant no. SP/19/3/34678). This study used anonymised data held in NHS England’s Secure Data Environment service for England. It is made available via the BHF Data Science Centre’s CVD-COVID-UK/COVID-IMPACT consortium. This work used data provided by patients and collected by the NHS as part of their care and support. We would like to acknowledge all data providers who make health-relevant data available for research. We also thank Huayu Zhang for his contributions to the foundational phenotyping workflows. This research is part of the Data and Connectivity National Core Study, led by Health Data Research UK in partnership with the Office for National Statistics and funded by UK Research and Innovation (grant ref MC_PC_20058). The Alan Turing Institute also supported this work via ‘Towards Turing 2.0’ EPSRC Grant Funding. The funders had no role in the study design, data collection, data analysis, data interpretation, or report writing.

## Reference

1. Joppi R, Bertele’ V, Garattini S. Orphan drugs, orphan diseases. The first decade of orphan drug legislation in the EU. Eur J Clin Pharmacol. 2013;69:1009–24. 10.1007/s00228-012-1423-2.

2. eClinicalMedicine. Raising the voice for rare diseases: under the spotlight for equity. eClinicalMedicine. 2023;57. 10.1016/j.eclinm.2023.101941.

3. Department of Health & Social Care. England Rare Diseases Action Plan 2025: main report. https://www.gov.uk/government/publications/england-rare-diseases-action-plan-2025/england-rare-diseases-action-plan-2025-main-report. Accessed 4 Aug 2025.

4. Davies S. Annual report of the Chief Medical Officer 2016, generation genome. Department of Health. 2017.

5. Bauskis A, Strange C, Molster C, Fisher C. The diagnostic odyssey: insights from parents of children living with an undiagnosed condition. Orphanet J Rare Dis. 2022;17:233. 10.1186/s13023-022-02358-x.

6. Adachi T, El-Hattab AW, Jain R, Nogales Crespo KA, Quirland Lazo CI, Scarpa M, et al. Enhancing Equitable Access to Rare Disease Diagnosis and Treatment around the World: A Review of Evidence, Policies, and Challenges. International Journal of Environmental Research and Public Health. 2023;20:4732. 10.3390/ijerph20064732.

7. Briscoe S, Martin Pintado C, Sutcliffe K, Melendez-Torres GJ, Garside R, Lawal HM, et al. Evidence of inequities experienced by the rare disease community with respect to receipt of a diagnosis and access to services: a scoping review of UK and international evidence. Orphanet Journal of Rare Diseases. 2025;20:303. 10.1186/s13023-025-03818-w.

8. Kapadia D, Zhang J, Salway S, Nazroo J, Booth A. Ethnic Inequalities in Healthcare: A Rapid Evidence Review.

9. Wojcik MH, Bresnahan M, del Rosario MC, Ojeda MM, Kritzer A, Fraiman YS. Rare diseases, common barriers: disparities in pediatric clinical genetics outcomes. Pediatr Res. 2023;93:110–7. 10.1038/s41390-022-02240-3.

10. Sirugo G, Williams SM, Tishkoff SA. The Missing Diversity in Human Genetic Studies. Cell. 2019;177:26–31. 10.1016/j.cell.2019.02.048.

11. Kelsey MD, Patrick-Lake B, Abdulai R, Broedl UC, Brown A, Cohn E, et al. Inclusion and diversity in clinical trials: Actionable steps to drive lasting change. Contemporary Clinical Trials. 2022;116:106740. 10.1016/j.cct.2022.106740.

12. Thygesen JH, Zhang H, Issa H, Wu J, Hama T, Phiho-Gomes A-C, et al. Prevalence and demographics of 331 rare diseases and associated COVID-19-related mortality among 58 million individuals: a nationwide retrospective observational study. Lancet Digit Health. 2025;7:e145–56. 10.1016/S2589-7500(24)00253-X.

13. Pineda-Moncusí M, Allery F, Delmestri A, Bolton T, Nolan J, Thygesen JH, et al. Ethnicity data resource in population-wide health records: completeness, coverage and granularity of diversity. Sci Data. 2024;11:221. 10.1038/s41597-024-02958-1.

14. Department of Health & Social Care. The UK Rare Diseases Framework. https://www.gov.uk/government/publications/uk-rare-diseases-framework/the-uk-rare-diseases-framework. Accessed 4 Aug 2025.

15. Equity in Health Research. Genomics England. https://www.genomicsengland.co.uk/initiatives/diverse-data/equity-in-health-research. Accessed 4 Aug 2025.

16. Office for National Statistics. Population and household estimates, England and Wales: Census 2021. https://www.ons.gov.uk/peoplepopulationandcommunity/populationandmigration/populationestimates/datasets/populationandhouseholdestimatesenglandandwalescensus2021. Accessed 23 June 2025.

17. Thygesen JH, Tomlinson C, Hollings S, Mizani MA, Handy A, Akbari A, et al. COVID-19 trajectories among 57 million adults in England: a cohort study using electronic health records. The Lancet Digital Health. 2022;4:e542–57. 10.1016/S2589-7500(22)00091-7.

18. Pineda-Moncusí M, Allery F, Abbasizanjani H, Powell D, Prats-Uribe A, Thygesen JH, et al. Ethnic disparities in COVID-19 mortality and cardiovascular disease in England and Wales between 2020-2022. Nat Commun. 2025;16:6059. 10.1038/s41467-025-59951-4.

19. Morales DR, Ali SN. COVID-19 and disparities affecting ethnic minorities. The Lancet. 2021;397:1684–5. 10.1016/S0140-6736(21)00949-1.

20. Tai DBG, Shah A, Doubeni CA, Sia IG, Wieland ML. The Disproportionate Impact of COVID-19 on Racial and Ethnic Minorities in the United States. Clin Infect Dis. 2021;72:703–6. 10.1093/cid/ciaa815.

21. Tai DBG, Sia IG, Doubeni CA, Wieland ML. Disproportionate Impact of COVID-19 on Racial and Ethnic Minority Groups in the United States: a 2021 Update. J Racial and Ethnic Health Disparities. 2022;9:2334–9. 10.1007/s40615-021-01170-w.

22. Wu M, Davis JD, Zhao C, Daley T, Oliver KE. Racial inequities and rare *CFTR* variants: Impact on cystic fibrosis diagnosis and treatment. Journal of Clinical & Translational Endocrinology. 2024;36:100344. 10.1016/j.jcte.2024.100344.

23. Newborn Genomes Programme. Genomics England. https://www.genomicsengland.co.uk/initiatives/newborns. Accessed 6 Aug 2025.

24. COVID-19: general public testing behaviours. GOV.UK. https://www.gov.uk/government/publications/lfd-tests-how-and-why-they-were-used-during-the-pandemic/covid-19-general-public-testing-behaviours. Accessed 11 Jan 2026.

