## Supplement for "Prevalence of 406 rare diseases by ethnicity and their associated COVID-19 infection burden: A national cross-sectional study of 62.5 million people in England"

### Supplementary tables

#### Supplementary table 1. Baseline Demographic Characteristics of the Study Cohort: By Rare Disease Categories

| **Characteristic** | **Cardiovascular**  **N = 198,265** | **Metabolic**  **N = 10,440** | **Other**  **N = 818,520** | **Mix**  **N = 24,245** | **P-value** |
| --- | --- | --- | --- | --- | --- |
| Age (years) |  |  |  |  |  |
| Age at index date | 26.0 (10.0-53.0) | 19.0 (15.0-52.0) | 53.0 (23.0-75.0) | 10.0 (5.0-18.0) | <0.001 |
| Age at diagnosis | 4.0 (0.0-38.0) | 3.0 (0.0-41.0) | 42.0 (9.0-68.0) | 0.0 (0.0-1.0) | <0.001 |
| Sex |  |  |  |  |  |
| Male | 100,800 (50.8%) | 5,125 (49.1%) | 357,070 (43.6%) | 12,860 (53.0%) | <0.001 |
| Female | 97,450 (49.2%) | 5,315 (50.9%) | 461,400 (56.4%) | 11,385 (47.0%) | |
| Unknown | 15 (0.0%) | <10 | 50 (0.0%) | NA |  |
| IMD quintile |  |  |  |  |  |
| 1 (Most deprived) | 45,790 (23.1%) | 2,330 (22.3%) | 166,900 (20.4%) | 6,545 (27.0%) | <0.001 |
| 2 | 40,430 (20.4%) | 1,890 (18.1%) | 160,415 (19.6%) | 5,395 (22.3%) | |
| 3 | 38,180 (19.3%) | 1,900 (18.2%) | 166,110 (20.3%) | 4,565 (18.8%) | |
| 4 | 37,100 (18.7%) | 1,840 (17.6%) | 165,280 (20.2%) | 4,025 (16.6%) | |
| 5 (Least deprived) | 36,615 (18.5%) | 2,470 (23.7%) | 159,285 (19.5%) | 3,710 (15.3%) | |
| Unknown | 145 (0.1%) | <10 | 540 (0.1%) | 10 (0.0%) |  |
| Number of rare diseases per patient | 1.0 (1.0-1.0) | 1.0 (1.0-1.0) | 1.0 (1.0-1.0) | 2.0 (2.0-3.0) | <0.001 |
| COVID-19 infection | 64,855 (32.7%) | 3,845 (36.8%) | 268,080 (32.8%) | 7,625 (31.4%) | <0.001 |
| ONS Ethnicity |  |  |  |  |  |
| White | 151,675 (76.5%) | 7,825 (75.0%) | 673,500 (82.3%) | 17,295 (71.3%) | <0.001 |
| Asian or Asian British | 17,985 (9.1%) | 1,285 (12.3%) | 50,780 (6.2%) | 2,655 (11.0%) | |
| Black or Black British | 7,165 (3.6%) | 195 (1.9%) | 22,835 (2.8%) | 1,195 (4.9%) | |
| Mixed | 5,025 (2.5%) | 200 (1.9%) | 14,170 (1.7%) | 970 (4.0%) |  |
| Other Ethnic Group | 3,475 (1.8%) | 160 (1.5%) | 9,480 (1.2%) | 535 (2.2%) |  |
| Unknown | 12,940 (6.5%) | 780 (7.5%) | 47,750 (5.8%) | 1,600 (6.6%) | |
| Primary Code Ethnicity |  |  |  |  |  |
| White British | 136,770 (69.0%) | 6,880 (65.9%) | 619,940 (75.7%) | 15,770 (65.0%) | <0.001 |
| White Irish | 1,050 (0.5%) | 45 (0.4%) | 5,175 (0.6%) | 90 (0.4%) |  |
| Gypsy or Irish Traveller | 165 (0.1%) | 15 (0.1%) | 385 (0.0%) | 20 (0.1%) |  |
| Any other White background | 13,695 (6.9%) | 885 (8.5%) | 48,000 (5.9%) | 1,410 (5.8%) | |
| Indian | 4,935 (2.5%) | 205 (2.0%) | 15,740 (1.9%) | 575 (2.4%) |  |
| Pakistani | 7,020 (3.5%) | 770 (7.4%) | 18,375 (2.2%) | 1,200 (4.9%) | |
| Bangladeshi | 2,175 (1.1%) | 85 (0.8%) | 5,165 (0.6%) | 340 (1.4%) |  |
| Chinese | 620 (0.3%) | 35 (0.3%) | 2,155 (0.3%) | 60 (0.2%) |  |
| Other Asian | 3,235 (1.6%) | 185 (1.8%) | 9,350 (1.1%) | 480 (2.0%) |  |
| Black African | 4,140 (2.1%) | 120 (1.1%) | 11,285 (1.4%) | 715 (2.9%) |  |
| Black Caribbean | 1,555 (0.8%) | 30 (0.3%) | 6,890 (0.8%) | 215 (0.9%) |  |
| Other Black | 1,465 (0.7%) | 45 (0.4%) | 4,655 (0.6%) | 270 (1.1%) |  |
| White and Asian | 1,050 (0.5%) | 40 (0.4%) | 2,720 (0.3%) | 185 (0.8%) |  |
| White and African | 775 (0.4%) | 30 (0.3%) | 2,245 (0.3%) | 145 (0.6%) |  |
| White and Caribbean | 1,360 (0.7%) | 50 (0.5%) | 4,010 (0.5%) | 240 (1.0%) |  |
| Arab | 350 (0.2%) | 10 (0.1%) | 850 (0.1%) | 55 (0.2%) |  |
| Other Ethnic Group | 3,120 (1.6%) | 150 (1.4%) | 8,630 (1.1%) | 475 (2.0%) |  |
| Other Mixed | 1,840 (0.9%) | 80 (0.8%) | 5,195 (0.6%) | 400 (1.6%) |  |
| Unknown | 12,940 (6.5%) | 780 (7.5%) | 47,750 (5.8%) | 1,600 (6.6%) |  |

Note: Data are presented as counts (percentages) for categorical variables or median (interquartile range) for continuous variables. Statistical tests used: Mann-Whitney U test for continuous variables and Chi-square test for categorical variables. In adherence with NHS England's data protection policies, any patient count below 10 was suppressed and displayed as "<10", and all other counts were rounded to the nearest five. Each of the 406 selected rare diseases was assigned by a clinician to one of three high-level categories based on its primary physiological system: ‘Cardiovascular’, ‘Metabolic’, or ‘Other’. At the patient level, individuals were assigned a patient-level category. If a patient had multiple rare diseases that all belonged to the same category, they were assigned that category. If a patient had diagnoses for rare diseases spanning more than one of these categories, they were classified as ‘Mixed’. This classification was used to describe the baseline characteristics of the rare disease cohort.

#### Supplementary table 2. Baseline Demographic Characteristics of the Study Cohort: By the Six Broad ONS Ethnicity Groups

| **Characteristic** | **White**  **N = 850,290** | **Asian or Asian British**  **N = 72,705** | **Black or Black British**  **N = 31,390** | **Mixed**  **N = 20,365** | **Other Ethnic Group**  **N =13,645** | **Unknown**  **N = 63,070** | **P-value** |
| --- | --- | --- | --- | --- | --- | --- | --- |
| Age (years): |  |  |  |  |  |  |  |
| Age at index date | 52.0 (21.0-74.0) | 24.0 (10.0-51.0) | 25.0 (11.0-54.0) | 15.0 (7.0-34.0) | 22.0 (8.0-49.0) | 30.0 (16.0-62.0) | <0.001 |
| Age at diagnosis | 39.0 (5.0-66.0) | 10.0 (0.0-41.0) | 14.0 (0.0-43.0) | 2.0 (0.0-22.0) | 9.0 (0.0-40.0) | 16.0 (1.0-53.0) | <0.001 |
| Sex |  |  |  |  |  |  |  |
| Male | 376,300 (44.3%) | 35,670 (49.1%) | 14,675 (46.8%) | 10,165 (49.9%) | 6,845 (50.2%) | 32,195 (51.0%) | <0.001 |
| Female | 473,935 (55.7%) | 37,035 (50.9%) | 16,715 (53.2%) | 10,200 (50.1%) | 6,800 (49.8%) | 30,870 (48.9%) | |
| Unknown | 60 (0.0%) | NA | <10 | NA | NA | <10 |  |
| IMD quintile |  |  |  |  |  |  |  |
| 1 (Most deprived) | 162,220 (19.1%) | 25,335 (34.8%) | 12,385 (39.5%) | 5,835 (28.7%) | 4,200 (30.8%) | 11,590 (18.4%) | <0.001 |
| 2 | 159,365 (18.7%) | 18,655 (25.7%) | 9,785 (31.2%) | 4,910 (24.1%) | 3,665 (26.9%) | 11,750 (18.6%) | |
| 3 | 174,800 (20.6%) | 12,205 (16.8%) | 4,910 (15.6%) | 3,755 (18.4%) | 2,440 (17.9%) | 12,645 (20.0%) | |
| 4 | 178,705 (21.0%) | 8,920 (12.3%) | 2,665 (8.5%) | 3,045 (15.0%) | 1,855 (13.6%) | 13,055 (20.7%) | |
| 5 (Least deprived) | 174,600 (20.5%) | 7,575 (10.4%) | 1,625 (5.2%) | 2,810 (13.8%) | 1,480 (10.8%) | 13,990 (22.2%) | |
| Unknown | 600 (0.1%) | 15 (0.0%) | 20 (0.1%) | 10 (0.0%) | <10 | 45 (0.1%) |  |
| Number of rare diseases per patient | 1.0 (1.0-1.0) | 1.0 (1.0-1.0) | 1.0 (1.0-1.0) | 1.0 (1.0-1.0) | 1.0 (1.0-1.0) | 1.0 (1.0-1.0) | <0.001 |
| COVID-19 infection | 285,100 (33.5%) | 21,390 (29.4%) | 8,250 (26.3%) | 5,995 (29.4%) | 3,640 (26.7%) | 20,025 (31.7%) | <0.001 |
| Rare Disease Category |  |  |  |  |  |  |  |
| Cardiovascular | 151,675 (17.8%) | 17,985 (24.7%) | 7,165 (22.8%) | 5,025 (24.7%) | 3,475 (25.5%) | 12,940 (20.5%) | <0.001 |
| Metabolic | 7,825 (0.9%) | 1,285 (1.8%) | 195 (0.6%) | 200 (1.0%) | 160 (1.2%) | 780 (1.2%) |  |
| Other | 673,500 (79.2%) | 50,780 (69.8%) | 22,835 (72.7%) | 14,170 (69.6%) | 9,480 (69.5%) | 47,750 (75.7%) | |
| Mix | 17,295 (2.0%) | 2,655 (3.7%) | 1,195 (3.8%) | 970 (4.8%) | 535 (3.9%) | 1,600 (2.5%) | |

Note: Data are presented as counts (percentages) for categorical variables or median (interquartile range) for continuous variables. Statistical tests used: Mann-Whitney U test for continuous variables and Chi-square test for categorical variables. In adherence with NHS England's data protection policies, any patient count below 10 was suppressed and displayed as "<10", and all other counts were rounded to the nearest five.

#### Supplementary table 3. Baseline Demographic Characteristics of the Study Cohort: By the Nineteen NHS Primary Care Ethnicity Categories

| **Characteristic** | **White British**  **N = 779,360** | **White Irish**  **N = 6,365** | **Gypsy or Irish Traveller**  **N = 580** | **Any other White background**  **N = 63,985** | **Indian**  **N = 21,455** | **Pakistani**  **N = 27,370** | **Bangladeshi**  **N = 7,765** | **Chinese**  **N = 2,870** | **Other Asian**  **N = 13,245** | **Black African**  **N = 16,260** | **Black Caribbean**  **N = 8,695** | **Other Black**  **N = 6,435** | **White and Asian**  **N = 3,990** | **White and African**  **N = 3,195** | **White and Caribbean**  **N = 5,665** | **Arab**  **N = 1,265** | **Other Ethnic Group**  **N = 12,375** | **Unknown**  **N = 63,070** | **Other Mixed**  **N = 7,515** | **P-value** |
| --- | --- | --- | --- | --- | --- | --- | --- | --- | --- | --- | --- | --- | --- | --- | --- | --- | --- | --- | --- | --- |
| Age (years): |  |  |  |  |  |  |  |  |  |  |  |  |  |  |  |  |  |  |  |  |
| Age at index date | 52.0 (22.0-75.0) | 67.0 (42.0-80.0) | 23.0 (8.0-49.0) | 43.0 (16.0-69.0) | 39.0 (13.0-63.0) | 19.0 (9.0-40.0) | 18.0 (8.0-38.0) | 34.0 (13.0-58.0) | 26.0 (10.0-53.0) | 19.0 (9.0-45.0) | 47.0 (20.0-64.0) | 22.0 (10.0-48.0) | 14.0 (7.0-29.0) | 15.0 (7.0-37.0) | 17.0 (7.0-35.0) | 17.0 (7.0-41.0) | 23.0 (9.0-50.0) | 30.0 (16.0-62.0) | 14.0 (6.0-34.0) | <0.001 |
| Age at diagnosis | 40.0 (5.0-66.0) | 58.0 (31.0-73.0) | 8.0 (0.0-40.0) | 33.0 (3.0-60.0) | 27.0 (1.0-53.0) | 4.0 (0.0-29.0) | 4.0 (0.0-29.0) | 23.0 (1.0-49.0) | 14.0 (0.0-44.0) | 6.0 (0.0-36.0) | 35.0 (4.0-55.0) | 9.0 (0.0-37.0) | 1.0 (0.0-16.0) | 3.0 (0.0-27.0) | 2.0 (0.0-24.0) | 7.0 (0.0-33.0) | 10.0 (0.0-41.0) | 16.0 (1.0-53.0) | 1.0 (0.0-23.0) | <0.001 |
| Sex |  |  |  |  |  |  |  |  |  |  |  |  |  |  |  |  |  |  |  |  |
| Male | 345,255 (44.3%) | 2,590 (40.7%) | 295 (50.8%) | 28,165 (44.0%) | 10,295 (48.0%) | 13,820 (50.5%) | 3,885 (50.0%) | 1,290 (45.0%) | 6,380 (48.2%) | 8,000 (49.2%) | 3,560 (41.0%) | 3,115 (48.4%) | 2,080 (52.1%) | 1,585 (49.6%) | 2,780 (49.1%) | 695 (54.9%) | 6,150 (49.7%) | 32,195 (51.0%) | 3,720 (49.5%) | <0.001 |
| Female | 434,050 (55.7%) | 3,775 (59.3%) | 285 (49.1%) | 35,820 (56.0%) | 11,160 (52.0%) | 13,550 (49.5%) | 3,880 (50.0%) | 1,575 (54.9%) | 6,865 (51.8%) | 8,260 (50.8%) | 5,130 (59.0%) | 3,325 (51.7%) | 1,910 (47.9%) | 1,610 (50.4%) | 2,885 (50.9%) | 570 (45.0%) | 6,225 (50.3%) | 30,870 (48.9%) | 3,795 (50.5%) |  |
| Unknown | 55 (0.0%) | NA | NA | <10 | NA | NA | NA | NA | NA | NA | <10 | NA | NA | NA | NA | NA | NA | <10 | NA |  |
| IMD quintile |  |  |  |  |  |  |  |  |  |  |  |  |  |  |  |  |  |  |  |  |
| 1 (Most deprived) | 147,945 (19.0%) | 1,165 (18.3%) | 180 (31.0%) | 12,930 (20.2%) | 3,825 (17.8%) | 14,075 (51.4%) | 3,470 (44.7%) | 525 (18.3%) | 3,440 (26.0%) | 6,800 (41.8%) | 3,130 (36.0%) | 2,455 (38.1%) | 900 (22.6%) | 990 (31.0%) | 1,960 (34.6%) | 480 (37.9%) | 3,720 (30.1%) | 11,590 (18.4%) | 1,985 (26.4%) | <0.001 |
| 2 | 144,000 (18.5%) | 1,445 (22.7%) | 120 (20.7%) | 13,800 (21.6%) | 5,355 (25.0%) | 6,665 (24.3%) | 2,530 (32.6%) | 605 (21.1%) | 3,500 (26.4%) | 4,975 (30.6%) | 2,830 (32.6%) | 1,975 (30.7%) | 760 (19.0%) | 860 (26.9%) | 1,435 (25.3%) | 320 (25.3%) | 3,340 (27.0%) | 11,750 (18.6%) | 1,855 (24.7%) |  |
| 3 | 160,065 (20.5%) | 1,305 (20.5%) | 135 (23.2%) | 13,290 (20.8%) | 4,670 (21.8%) | 3,245 (11.9%) | 990 (12.7%) | 560 (19.5%) | 2,740 (20.7%) | 2,320 (14.3%) | 1,540 (17.7%) | 1,050 (16.3%) | 755 (18.9%) | 545 (17.0%) | 1,025 (18.1%) | 230 (18.2%) | 2,205 (17.8%) | 12,645 (20.0%) | 1,430 (19.0%) |  |
| 4 | 165,010 (21.2%) | 1,265 (19.9%) | 85 (14.6%) | 12,345 (19.3%) | 3,895 (18.2%) | 2,015 (7.4%) | 490 (6.3%) | 555 (19.4%) | 1,965 (14.8%) | 1,310 (8.1%) | 750 (8.6%) | 605 (9.4%) | 735 (18.4%) | 450 (14.1%) | 695 (12.3%) | 155 (12.2%) | 1,705 (13.8%) | 13,055 (20.7%) | 1,160 (15.4%) |  |
| 5 (Least deprived) | 161,790 (20.8%) | 1,180 (18.5%) | 60 (10.3%) | 11,565 (18.1%) | 3,705 (17.3%) | 1,365 (5.0%) | 280 (3.6%) | 625 (21.8%) | 1,600 (12.1%) | 840 (5.2%) | 435 (5.0%) | 350 (5.4%) | 840 (21.1%) | 350 (10.9%) | 545 (9.6%) | 80 (6.3%) | 1,395 (11.3%) | 13,990 (22.2%) | 1,080 (14.4%) |  |
| Unknown | 550 (0.1%) | <10 | NA | 50 (0.1%) | <10 | <10 | <10 | <10 | <10 | 10 (0.1%) | <10 | <10 | <10 | <10 | <10 | NA | <10 | 45 (0.1%) | <10 |  |
| Number of rare diseases per patient | 1.0 (1.0-1.0) | 1.0 (1.0-1.0) | 1.0 (1.0-1.0) | 1.0 (1.0-1.0) | 1.0 (1.0-1.0) | 1.0 (1.0-1.0) | 1.0 (1.0-1.0) | 1.0 (1.0-1.0) | 1.0 (1.0-1.0) | 1.0 (1.0-1.0) | 1.0 (1.0-1.0) | 1.0 (1.0-1.0) | 1.0 (1.0-1.0) | 1.0 (1.0-1.0) | 1.0 (1.0-1.0) | 1.0 (1.0-1.0) | 1.0 (1.0-1.0) | 1.0 (1.0-1.0) | 1.0 (1.0-1.0) | <0.001 |
| COVID-19 infection | 262,860 (33.7%) | 1,900 (29.9%) | 140 (24.1%) | 20,205 (31.6%) | 6,865 (32.0%) | 7,590 (27.7%) | 2,115 (27.2%) | 815 (28.4%) | 4,010 (30.3%) | 3,865 (23.8%) | 2,615 (30.1%) | 1,770 (27.5%) | 1,215 (30.5%) | 890 (27.8%) | 1,730 (30.5%) | 305 (24.1%) | 3,335 (26.9%) | 20,025 (31.7%) | 2,160 (28.7%) | <0.001 |
| Rare Disease Category |  |  |  |  |  |  |  |  |  |  |  |  |  |  |  |  |  |  |  |  |
| Cardiovascular | 136,770 (17.5%) | 1,050 (16.5%) | 165 (28.4%) | 13,695 (21.4%) | 4,935 (23.0%) | 7,020 (25.6%) | 2,175 (28.0%) | 620 (21.6%) | 3,235 (24.4%) | 4,140 (25.5%) | 1,555 (17.9%) | 1,465 (22.8%) | 1,050 (26.3%) | 775 (24.2%) | 1,360 (24.0%) | 350 (27.6%) | 3,120 (25.2%) | 12,940 (20.5%) | 1,840 (24.5%) | <0.001 |
| Metabolic | 6,880 (0.9%) | 45 (0.7%) | 15 (2.6%) | 885 (1.4%) | 205 (1.0%) | 770 (2.8%) | 85 (1.1%) | 35 (1.2%) | 185 (1.4%) | 120 (0.7%) | 30 (0.3%) | 45 (0.7%) | 40 (1.0%) | 30 (0.9%) | 50 (0.9%) | 10 (0.8%) | 150 (1.2%) | 780 (1.2%) | 80 (1.1%) |  |
| Other | 619,940 (79.5%) | 5,175 (81.3%) | 385 (66.3%) | 48,000 (75.0%) | 15,740 (73.4%) | 18,375 (67.1%) | 5,165 (66.5%) | 2,155 (75.1%) | 9,350 (70.6%) | 11,285 (69.4%) | 6,890 (79.3%) | 4,655 (72.3%) | 2,720 (68.2%) | 2,245 (70.2%) | 4,010 (70.8%) | 850 (67.1%) | 8,630 (69.7%) | 47,750 (75.7%) | 5,195 (69.1%) |  |
| Mix | 15,770 (2.0%) | 90 (1.4%) | 20 (3.4%) | 1,410 (2.2%) | 575 (2.7%) | 1,200 (4.4%) | 340 (4.4%) | 60 (2.1%) | 480 (3.6%) | 715 (4.4%) | 215 (2.5%) | 270 (4.2%) | 185 (4.6%) | 145 (4.5%) | 240 (4.2%) | 55 (4.3%) | 475 (3.8%) | 1,600 (2.5%) | 400 (5.3%) |  |

Note: Data are presented as counts (percentages) for categorical variables or median (interquartile range) for continuous variables. Statistical tests used: Mann-Whitney U test for continuous variables and Chi-square test for categorical variables. In adherence with NHS England's data protection policies, any patient count below 10 was suppressed and displayed as "<10", and all other counts were rounded to the nearest five.

### Supplementary Figures

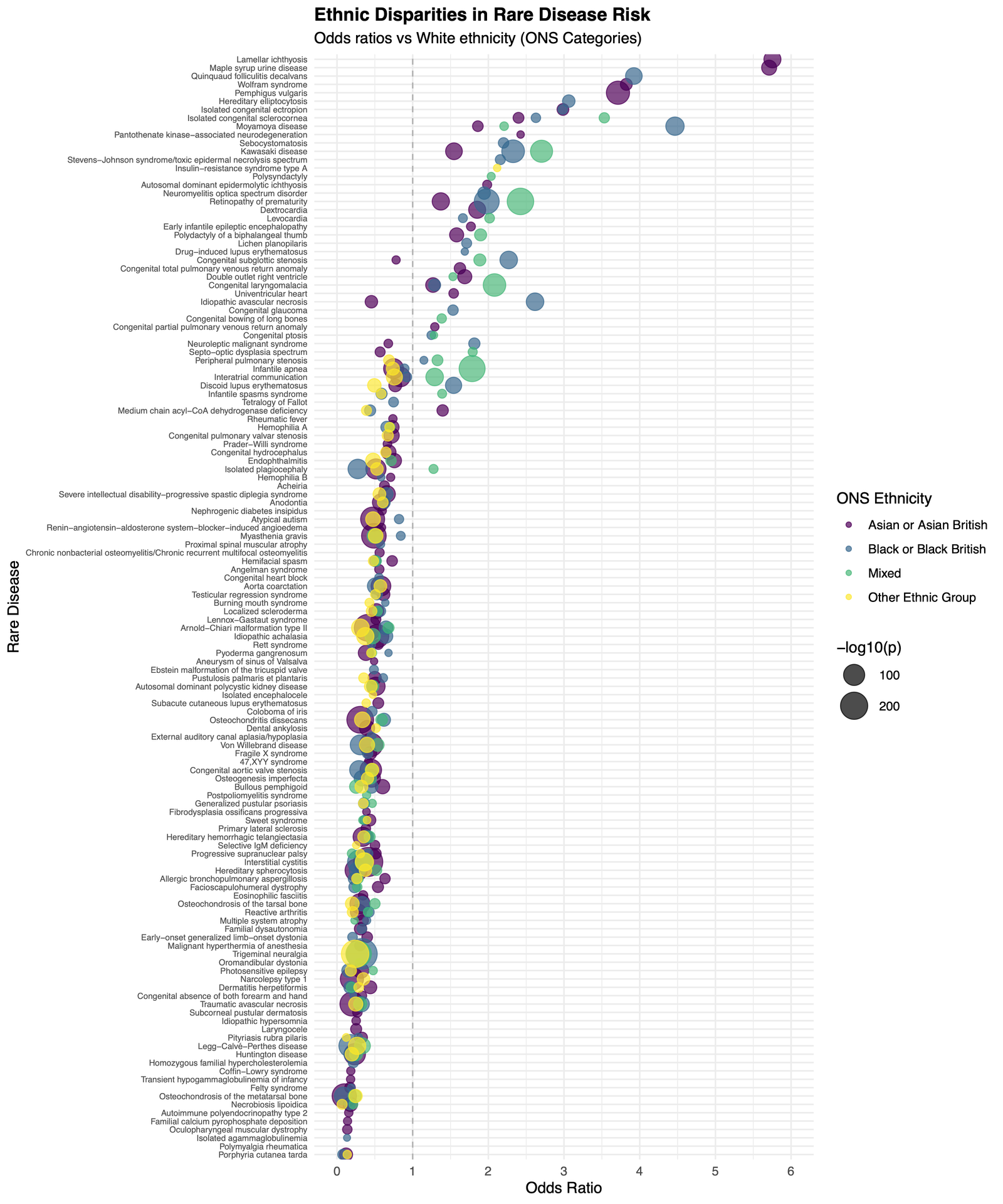

#### Supplementary figure 1. Ethnic Disparities in Rare Disease Risk: Odds Ratios Relative to White Ethnicity

Bubble plot depicting odds ratios for the risk of various rare diseases across ethnic groups, using White ethnicity as the reference category. Each point represents a rare disease, stratified by Office for National Statistics (ONS) ethnicity categories: Asian or Asian British (purple), Black or Black British (blue), Mixed (green), and Other Ethnic Group (yellow). The size of each bubble is proportional to the statistical significance (−log10(p-value)) of the association. The x-axis indicates the odds ratio, while the y-axis lists rare diseases.

### Supplementary Data

#### Supplementary data 1. Rare disease categories

Each of the 406 selected rare diseases was assigned by a clinician to one of three high-level categories based on its primary physiological system: ‘Cardiovascular’, ‘Metabolic’, or ‘Other’. At the patient level, individuals were assigned a patient-level category. If a patient had multiple rare diseases that all belonged to the same category, they were assigned that category. If a patient had diagnoses for rare diseases spanning more than one of these categories, they were classified as ‘Mixed’. This classification was used to describe the baseline characteristics of the rare disease cohort.

#### Supplementary data 2. The age- and sex-standardised prevalence for 406 rare diseases.

Point prevalence per million individuals for each of the 406 rare diseases was calculated as the number of individuals with a diagnosis alive on the index date. These crude rates were then adjusted for age and sex using the direct standardisation method, with the 2021 England Census population serving as the reference standard population[15]. 95% confidence intervals (CIs) for the adjusted prevalence rates were calculated based on the variance of the standardised rates.

#### Supplementary data 3. Prevalence of rare diseases by ethnicity: ONS high-level and NHS primary care categories

Point prevalence per million individuals for each of the 406 rare diseases by ethnicity. In adherence with NHS England's data protection policies, any patient count below 10 was suppressed and displayed as "<10", and all other counts were rounded to the nearest five.

#### Supplementary data 4. Ethnicity differences (ONS high-level categories)

Test statistics for Fishers exact analysis of six ONS high-level ethnicity differences of the 227 rare diseases with

100 or more patients identified. Ethnicity analysis compares Asian or Asian British, Black or Black British, Mixed, and Other Ethnic Group to the majority White ethnicity. In adherence with NHS England's data protection policies, any patient count below 10 was suppressed and displayed as "<10", and all other counts were rounded to the nearest five.

#### Supplementary data 5. Ethnicity differences (NHS primary care categories)

Test statistics for Fishers exact analysis of nineteen NHS primary care ethnicity differences of the 227 rare diseases with 100 or more patients identified. Ethnicity analysis compares White Irish, Gypsy or Irish Traveller, Any other White background, Indian, Pakistani, Bangladeshi, Chinese, Other Asian, Black African, Black Caribbean, Other Black, White and Asian, White and African, White and Caribbean, Arab, Other Ethnic Group, Other Mixed to the majority White British ethnicity. In adherence with NHS England's data protection policies, any patient count below 10 was suppressed and displayed as "<10", and all other counts were rounded to the nearest five.

#### Supplementary data 6. Gender differences

Test statistics for Fishers exact analysis of gender differences of the 227 rare diseases with 100 or more patients identified. In adherence with NHS England's data protection policies, any patient count below 10 was suppressed and displayed as "<10", and all other counts were rounded to the nearest five.

#### Supplementary data 7. Percentage of patients with documented SARS-CoV-2 infection during the pandemic period.

COVID-19 infection status was defined as any documented positive test result (PCR or lateral flow assay) recorded between the start of the pandemic and the study index date (31 July 2023). For each of the 406 rare disease phenotypes, we calculated disease-specific COVID-19 infection rates as the proportion of patients with at least one documented positive test, expressed as percentages. In adherence with NHS England's data protection policies, any patient count below 10 was suppressed and displayed as "<10", and all other counts were rounded to the nearest five.
